# CVA-Flow novel telestroke system preliminary trial: reliability in determining NIHSS for acute ischemic stroke

**DOI:** 10.1101/2021.04.07.21254957

**Authors:** Saban Mor, Moskovitz Anner, Ohanian Sona, Reznik Anna, Ribo Marc, Sivan-Hoffmann Rotem

**Author notes:** **Correspondence author:** Mor saban; BA, MA, MPH, MEM, PhD, The Gertner Institute for Epidemiology and Health Policy Research, Ramat Gan, Israel. Equal contribution. The research has not been presented. All authors attest to meeting the ICMJE.org authorship criteria. Disclosure of Potential Conflicts of Interest and affiliation for all authors are attached separately. The authors received no specific funding for this work.

## Abstract

**Background:** The National Institutes of Health Stroke Scale (NIHSS) is the most recommended tool for objectively quantifying the impairment caused by a suspected stroke. Nevertheless, it is used almost solely by trained neurologists in the emergency departments (ED) setting. CVA-Flow (CVAid medical Ltd., Tel-Aviv, Israel) is a smartphone-based Telestroke system that captures the full NIHSS by video and enables a distant stroke physician to assess the patient’s neurological status, bringing forward the NIHSS to the pre-hospital setting.

**Objective:** We aimed to compare the reliability of an NIHSS score determined by a neurologist from afar, using the CVA-Flow platform, with a standard NIHSS assessment performed in the ED).

**Methods:** In this multi-center prospective trial, Patients admitted to the ED in Rambam hospital in Haifa, Israel, and Vall d’Hebron hospital (VdH) in Barcelona, Spain, had a neurological exam based on the NIHSS while being recorded by the system. A neurologist blinded to the results rated the NIHSS according to the videos offline.

**Results:** A total of 95 patients with a suspected stroke were included. Overall ICC was 0.936 (0.99 in VdH and 0.84 in Rambam), indicating excellent and good reliability, respectively.

**Conclusion:** Remote stroke assessment based on the NIHSS, using videos collected by the CVA-Flow platform, installed on a standard smartphone, is a reliable measurement as compared with bedside evaluation.

## Introduction

Management of acute ischemic stroke has changed dramatically following the evidence of the superiority of endovascular treatment (EVT) over best medical management, in the treatment of patients suffering large vessel occlusion (LVO), within 24 hours from onset of symptoms.^1–8^ The immediate consequence was an increase in the number of patients eligible for EVT, requiring secondary transfer of those patients from primary stroke centers to comprehensive stroke centers, capable of performing EVT.

EVT requires accurate and rapid diagnosis in the prehospital setting, as it is only indicated in specific patients with LVO, making up a small percentage of stroke cases,^9^ and its beneficial effects are highly time-dependent.^10^

Of the existing diagnostic tools that accurately distinguish LVO cases from non-LVO ones, the National Institutes of Health Stroke Scale (NIHSS) is the most recommended tool by healthcare providers to objectively quantify the neurological impairment caused by a suspected stroke.^11,12^ Nevertheless, the NIHSS is not considered to be feasible in the pre-hospital setting of EMS, since it requires a greater degree of training, is thought to be too time-consuming, and has not been as well validated in the prehospital setting.^12^

Several previous studies showed feasibility and reliability of the use of telemedicine for determining the NIHSS score from afar in stroke patients in real time,^13–21^ in simulated stroke patients in real time^22–26^ and captured video segments.^23–25^ Previous studies did not use designated applications designed for performing neurological evaluations, relying on simple applications for video conferencing instead, therefore requiring applicators to possess knowledge in performing neurological examinations and evaluators to be online the whole examination time.

To resolve this issue, CVAid Medical Ltd. developed a smartphone-based Telestroke system named CVA-Flow. This system aims to evaluate the patient’s neurological status, particularly detection of a possible stroke, assessment of stroke severity, and prediction of LVO as the cause of stroke. The system’s app guides the user through the examination step by step and captures the full NIHSS by short video segments, being transfer offline and enables a distant stroke physician to assess the patient’s status manually.

In this preliminary study, we aim to evaluate the reliability of an NIHSS score by a remote neurologist rating captured video examinations, using the CVA-Flow platform, compared to a standard NIHSS assessment done by a bedside neurologist in the ED.

## Methods

### Study Design and Participants

This multi-center study includes patients admitted to the ED in Rambam hospital in Haifa, Israel, and Vall d’Hebron hospital in Barcelona, Spain. Inclusion criteria were: (1) Age over 18 years of age, (2) Symptoms suggestive of stroke, (3) Had not received treatment (IVT or EVT) before the examination. Exclusion criteria were: (1) Patient intubated upon arrival, (2) Time from stroke symptoms onset greater than 24h, (3) Post-treatment (IVT or EVT), (4) Patient had other condition which could simulate signs of stroke (such as hypoglycemia).

Rambam hospital in Haifa serves as a comprehensive stroke center throughout the north of Israel and is a referral center for 12 district hospitals. Approximately 5,000 patients with symptoms suggestive of stroke are annually assessed by a neurologist in the hospital’s ED.

Vall d’Hebron hospital in Barcelona is one of the largest in Spain and serves as a comprehensive stroke center, which provides urgent stroke care to their area of reference and in other centers, that do not have a neurologist, via telemedicine. The ED of the general hospital screens annually about 3000 patients with symptoms suggestive of an acute stroke and admits about 1,500 patients.

Following an introduction by the researcher and signing an informed consent form, each subject was allocated a number by the application for anonymization. Following that, the subject was instructed by the researcher through a neurological exam based on the NIHSS with simple instructions presented on the screen, while being recorded by CVA-Collector. The application uses the smartphone’s built-in camera and microphone, to capture the clinical signs of the patient, and the data collected is uploaded in segments of the exam, using a secured network to the company’s server. The video is then sent in short segments representing each of the NIHSS tests to a remote physician, using CVA-Med, an application enabling the stroke team to watch the full exams from afar. An independent neurologist blinded to the score obtained on-site rated the NIHSS according to the videos remotely and offline (Figure 1). The results were compared to the NIHSS score assigned by one of five independent bedside neurologists from the relevant hospital as part of the routine clinical practice.

All raters are NIHSS-certified.

### Ethical Considerations

The study protocol was approved by the Institutional Human Subjects Ethics Committee (IRB-0187-18). All procedures performed were in accordance with the ethical standards of the institutional and national research committee. A detailed information fact in simple non-technical language was provided in advance and all patients eligible for signing included in the study were requested to sign an informed consent. Patients who were found ineligible of signing were included after receiving the family approval and signing of an independent physician. No compensation was provided for participating patients.

### Statistical methods

To validate the reliability of the collected data, the digitalized total score of the NIHSS was correlated with the results of the standard, bedside NIHSS examination.

Interrater reliability of the total NIHSS score was quantified using an intraclass correlation coefficient (ICC) model (2,1),^27^ namely two-way random effects, absolute agreement, and single rates/measurement. This method was previously used in corresponding articles.^20–26^ Based on the 95% confidence interval of the ICC estimate, ICC values of less than 0.5, 0.5 to 0.75, 0.75 to 0.9, and greater than 0.9 indicate poor, moderate, good, and excellent reliability, respectively.^28^

Interrater reliability of individual item scores was quantified using a conservative weighted Kappa (wK) coefficient. Kappa values of less than 0, 0 to 0.2, 0.2 to 0.4, 0.4 to 0.6, 0.6 to 0.8 and greater than 0.8 indicate no, slight, fair, moderate, substantial and perfect agreement, respectively.^29^

One sample Student’s t-test was used to determine whether the difference between total NIHSS scores differs from 0 significantly. Provided that the test was not significant, sensitivity to detect a change of the total NIHSS score was to be estimated at different levels of confidence using the Minimal Detectable Difference and Bland-Altman plots.^28,30^

We also examined if the difference between scores set from afar and from the bedside was over two NIHSS points by using a one-sample Student’s t-test. Following that, we calculated the ICC considering successful correlation if bedside rating was within 2 points of the remote rating.

The level of significance for all statistical analyses was 5%. The data analysis was performed using the Statistical Package for Health & Welfare Science for Windows (SPSS, version 25.0, Chicago, IL, USA).

### Sample size

The sample size was calculated by using Walter, Eliasziw & Donner (1998)^30^ web-based sample size calculator for reliability studies and statistical requirements for models of this type. Based on a medium effect size of 0.15, two repeated observations by different raters per subject, test power of 70%, significance of 0.05, acceptable reliability of 0.7, expected reliability of 0.8, there is a minimal need for 89 patients.

## Results

A total of 95 patients suffering from a suspected stroke were included, 44 from the ED in Rambam and 51 from the ED in Vall d’Hebron. Patient characteristics and results are summarized in Table 1. The total number of patients subsequently diagnosed with LVO was 32 (33.6%) with no differences in prevalence between centers. The percentage of males was slightly higher in Rambam compared with Vall d’Hebron. ICC (2,1) overall and in Vall d’Hebron was higher than 0.9, and for Rambam higher than 0.75, indicating excellent and good reliability, respectively.^28^

**Table 1:** Patient characteristics and intraclass correlation coefficient (ICC) of total National Institute of Health Stroke Scale score. Age is presented as mean ± standard deviation. Male and LVO rates are presented as absolute number and percentage. NIHSS is presented as median. IQR, Interquartile Range; LVO, Large Vessel Occlusion; N, number of patients; NIHSS, National Institute of Health Stroke Scale. *P-value < 0.001.

| Study site | Total | Rambam | Vall d'Hebron |
| --- | --- | --- | --- |
| N | 95 | 44 | 51 |
| Age | 69.93 ± 14.11 | 69.89 ± 13.77 | 69.06 ± 15.72 |
| Male | 50 (52.6%) | 27 (61.4%) | 23 (45.1%) |
| NIHSS | 5 [IQR, 5] | 5 [IQR, 6] | 4 [IQR, 7] |
| LVO | 32 (33.7%) | 15 (34.1%) | 17 (33.3%) |
| ICC (2,1) | 0.936* | 0.847* | 0.991* |

Subscale analysis for the individual reliability (wK) are summarized in Table 2. wK scores ranged from 0.285 to 0.646, indicating fair to substantial agreement, with the highest individual reliability observed in item 1b Level of Consciousness (LOC) Questions, followed by item 3, Visual and item 11, Extinction and Inattention. The lowest was in item 7, Limb Ataxia. The P-value for all Kappa scores was <0.001.

**Table 2:** Interrater agreement (weighted Kappa) on the subscales of the National Institute of Health Stroke Scale. CI, Confidence Interval; LOC, Level of Consciousness; wK, weighted Kappa. *p-Value < 0.001.

| Subscale | wK (95% CI; n=95) |
| --- | --- |
| 1a Level of Consciousness | 0.326* (0.045-0.607) |
| 1b LOC Questions | 0.646* (0.511-0.782) |
| 1c LOC Commands | 0.407* (0.162-0.652) |
| 2 Gaze | 0.567* (0.37-0.764) |
| 3 Visual | 0.633* (0.477-0.79) |
| 4 Facial Palsy | 0.461* (0.325-0.597) |
| 5a Motor Arm: Left Arm | 0.503* (0.355-0.65) |
| 5b Motor Arm: Right Arm | 0.355* (0.173-0.538) |
| 6a Motor Leg: Left Leg | 0.432* (0.278-0.586) |
| 6b Motor Leg: Right Leg | 0.398* (0.201-0.596) |
| 7 Limb Ataxia | 0.285* (0-0.57) |
| 8 Sensory | 0.491* (0.306-0.676) |
| 9 Language aphasia | 0.499* (0.339-0.658) |
| 10 Dysarthria | 0.458* (0.314-0.602) |
| 11 Extinction and Inattention | 0.626* (0.432-0.819) |

One sample Student’s t-test showed the difference between scores differs from 0 significantly (p < 0.05), therefor Minimal Detectable Difference and Bland–Altman plot were unnecessary.

One sample Student’s t-test showed the total NIHSS scores set from afar compared with those set at bedside did not differ significantly more than two points, (p = 0.325). We then calculated the ICC (2,1) adjusted for NIHSS scores with disagreement within two NIHSS points. The results indicate excellent reliability (ICC=0.939).

Intra-rater reliability for the independent remote neurologists who rated the NIHSS according to the offline videos and for the five bedside neurologists who rated the NIHSS as part of the routine clinical practice was high^18^ for both raters groups (Intraclass Correlation Coefficient: 0.89 and 0.87, respectively).

## Discussion

In this study we found the reliability of an NIHSS score determined by a neurologist from afar, using the CVA-Flow platform to be high compared to a standard NIHSS assessment done by a bedside neurologist in the ED.

Our results suggest that the total NIHSS determined by a neurologist using the system has a good to excellent reliability compared with a standard NIHSS assessment performed at the bedside. The use of recordings rather than real-time examination described in previous studies^13–26^ is an important part of the validation process of the CVA-Flow application and is the building block for the future artificial intelligence determination of stroke and LVO, yet to be installed. This artificial intelligence is designed to be trained on recordings and will perform the analysis after each segment is captured.

Interrater agreement on the total NIHSS score was consistent with preceding studies on real patients (0.91 - 0.98).^14,16–19^ Similar variability in the subscale items’ reliabilities was also seen.^13–19^ It should be noted that most previous studies were small.

The dissimilarities in the subscale analysis probably stem from the target study population characteristics and scale, exclusion criteria, and technological variations. Shafqat et al^13^ for example showed comparable results for all subscales except motor arm (5 a and b) and motor leg (6 a and b), which may be attributed to their use of a dual-camera system (wide-angle and zoom) in contrast to our use of smartphone camera.

The lowest reliability was observed in limb ataxia (7) as noted in many other studies using telemedicine on stroke patients (wK −0.07 to 0.35 in most studies).^13,14,16,18,19^

Differences between the centers are also an important finding. The bedside NIHSS was performed in Rambam by on-call, usually resident, neurologists. In Vall d’Hebron it was performed by neurologists from the stroke team. The NIHSS assessment from afar was done in both centers by skilled senior neurologists. Higher correlation in Vall d’Hebron suggests that the evaluation carried out through the system by a senior neurologist might be more accurate than the one performed by a resident, bedside. If this is true, using the system, both in ED, and even more in the pre-hospital setting, could increase the accuracy of NIHSS determination.

This study serves as a validation of the platform, indicating that NIHSS determination from afar on captured video segments using the dedicated application may be is as good as bedside assessment. Thus, enabling the possibility for future studies regarding assessment of stroke and LVO utilizing the CVA-Flow artificial intelligence assisted program and a standard smartphone, without the need for special equipment or hardware.

## Limitations

Limitation of this study is a lack of stroke mimics which might have decreased the variance and thus improved the results. Further research is needed for validation of the use of the system in such conditions. This study did not include a measurement of the time elapsed during an examination.

## Conclusions

Remote stroke assessment based on the NIHSS, using videos collected by the CVA-Flow platform installed on a standard smartphone, is significantly correlated with bedside evaluation, and therefore could be used as a reliable measurement for stroke and LVO prediction.

## Data Availability

The authors confirm that the data supporting the findings of this study are available within the article

